# Restoration of upper-extremity function after task-oriented, intention-driven functional electrical stimulation therapy using a wearable sleeve in adults with chronic stroke: a case series

**DOI:** 10.1101/2024.01.18.24301486

**Authors:** Ian W. Baumgart, Michael J. Darrow, Nicholas J. Tacca, Collin F. Dunlap, Samuel C. Colachis, Ashwini Kamath, Bryan R. Schlink, Philip T. Putnam, Joshua Branch, David A. Friedenberg, Lauren R. Wengerd, Eric C. Meyers

**Affiliations:** Neurotechnology, Battelle Memorial Institute; NeuroTech Institute, The Ohio State University

**Keywords:** Functional electrical stimulation therapy, intention-driven electrical stimulation, neuroplasticity, electromyography-driven electrical stimulation.

## Abstract

**Background:** Functional electrical stimulation (FES) has been recognized for decades as a method to retrain the motor system after stroke. Benefits of FES rehabilitation can be enhanced by combining task-oriented therapy, dubbed FES therapy (FEST). Furthermore, by synchronizing FES with the user’s volitional motor intention and incorporating multiple trained tasks FES can be better integrated into common task-oriented rehabilitation practice. Using wearable FES technology, we tested therapy incorporating these elements in two chronic stroke survivors.

**Methods:** Our group has developed the NeuroLife® Sleeve, a wearable forearm sleeve that contains a high-density grid of embedded FES electrodes, that may be controlled by an operator or by the wearer’s own electromyographic (EMG) signals. During eight weeks of FEST, intention-driven FES enabling multiple movements was delivered via operator control twice weekly and EMG control once weekly.

**Results:** At the end of the therapy period, subjects A and B had both improved their scores: Box and Blocks Test (A: +5, B: +7), the Action Arm Research Test (A: +7, B: +12), the Fugl Meyer Upper Extremity section (A: +11, B: +9), and the 9-Hole Peg Test (A: 158 sec, B: 54 sec, both previously unable). All score improvements persisted over the 10-week follow-up period despite greatly reduced (>80%) effective dose of FES.

**Conclusions:** This case series provides additional evidence that intention-driven FEST drives long-lasting motor recovery in chronic stroke survivors. The NeuroLife Sleeve enabled this therapy through the easily donned wearable sleeve interface, control schemes for pairing FES with motor intention, and efficient transitions between tasks with programmable FES placement and parameters.

## Introduction

Each year, nearly 800,000 people in the United States suffer a new or recurrent stroke^1^. Unilateral weakness, or hemiparesis, affects over 60% of stroke survivors^1^, and fewer than 20% of stroke survivors regain complete function in their affected arm despite months of rehabilitation^2^. These deficits in upper extremity function are profoundly disabling and significantly affect one’s ability to complete activities of daily living^3^. To reduce impairment after stroke, motor rehabilitation is the gold standard. However, impairment persists for many patients, making novel efficacious interventions an urgent priority.

Functional electrical stimulation (FES), in which electrical stimulation is used to evoke functional movement, is widely used as an adjuvant in stroke rehabilitation^4^. However, existing FES technologies offer limited opportunities to implement promising strategies to enhance the efficacy of FES. These strategies include combination with task-oriented training (TOT), synchronization of stimulation with motor intention, and practice spanning a range of movements and tasks. Below, we describe the evidence supporting each of the three strategies.

Task-oriented therapy, in which patients perform goal-directed functional task practice, generates greater neuroplastic effects compared to non-functional motor rehabilitation^5,6^. This type of training encourages active engagement from the patient, which is critical for driving neuroplasticity and optimal motor recovery^7,8^. Furthermore, successful completion of tasks provides reward feedback which drives motor skill learning and reorganization of neural circuitry to support long-lasting recovery^9^. Popovic and colleagues^10,11^ have defined the concept of FES therapy (FEST) as FES-enabled task training with volitional intent from the recipient and therapist assistance^10–13^. FEST delivered with pushbutton control was shown to be superior to conventional occupational therapy alone in 8-week courses of spinal cord injury and stroke upper limb rehabilitation^10,12,13^. These studies demonstrate that FES can be feasibly combined with TOT to improve the functional benefits of therapy, and motivates further study^14^.

While encouraging, these studies use stimulation with pre-determined duration initiated by user or therapist. This approach only requires motor intention to initiate stimulation but does not account for the user’s intention to sustain or end stimulation. Closer pairing of motor intention and FES is believed to enhance stroke recovery through principles of Hebbian neuroplasticity by synchronizing descending motor intent commands with the appropriate assisted movement and resultant ascending sensory activation^15,16^. EEG-based user control has been used to automatically pair motor intention with FES, generating upper limb functional improvement in chronic hemiplegic stroke survivors^17,18^. In these studies, EEG was used to initiate single movements or timed sequences, but more continuous user control over FES may confer a greater sense of agency, which has been suggested to support adaptive neuroplasticity and longer term “carryover effects” of FES^19^. Electromyography (EMG) has been investigated as an intuitive and more continuous control signal for intent-driven FES and FEST interventions^20–24^. EMG-controlled FES has been shown to drive corticospinal plasticity similarly to EEG-controlled^25^, however optimal parameters have yet to be identified and hardware limitations to use cases to current EMG-controlled FES systems limit conclusions regarding its relative efficacy^22^.

One such use case, using a variety of movements and tasks trained during FEST sessions, is limited by currently available technologies. Clinicians using TOT commonly tailor the tasks and difficulty to the patient’s ability and need, and practice multiple tasks each rehabilitation session^26^. However, most intention-driven FEST systems, such as FIT-FES, MyndMove and recoveriX, use adhesive patch electrodes requiring manual, trial-and-error placement by FES-trained therapists, leading to lost therapy time between tasks^13,23,27,28^. Wearable FEST systems, such as the Bioness H200, enable consistent electrode placement, but enable typically less than 4 functional movements^29^. An ideal FEST system will combine the flexibility to change stimulation locations and parameters quickly with the consistency of wearable electrode placement.

To solve these gaps in FEST technologies, our group has created the NeuroLife® Sleeve with two control schemes to pair FES with movement intention: operator- and EMG-controlled. The NeuroLife Sleeve enables a therapy program incorporating all of these discussed elements: (1) task-oriented training, (2) intention-driven assistance, (3) efficient switching between FES-enabled movements to facilitate multi-task practice during sessions. The NeuroLife Sleeve is a wearable forearm sleeve containing a high-definition array of up to 160 embedded electrodes that deliver individually programmable electrical stimulation. The garment was designed to allow for range of motion during FEST, and laptop graphical user interface (GUI) and EMG control schemes enable pairing with motor intention. Switching between stimulation configurations via GUI enables efficient transition between different grasps and functional movements without the need to reposition electrodes. In this case series, the NeuroLife Sleeve was controlled by operator and EMG schemes to deliver intention-driven FEST incorporating multiple movements. We demonstrate the capability to administer this intervention with the systems and report improved scores across multiple assessments of stroke recovery for both subjects.

## Materials and Methods

### Subjects

This study enrolled adults with upper limb hemiparesis, stroke-related hand impairment that interferes with the ability to complete activities of daily life, and who were classified as Stage 1-6 on the hand subscale of the Chedoke McMaster Stroke assessment. Individuals actively participating in stroke-related upper limb rehabilitation, co-occurring neurological or neuromuscular conditions, or implanted electronic devices were excluded. A complete list of inclusion and exclusion criteria are included in the supplementary materials.

### Materials

Two NeuroLife Sleeve systems were used to deliver FES throughout rehab. The EMG-controlled FES system (Fig. 1A) can both sense movement intent via EMG and stimulate via FES, whereas the FES-only system (Fig. 1B) delivers FES via a graphical user interface (GUI). Both systems consist of the fabric sleeve worn on the forearm, with a high-density array of up to 160 embedded stainless-steel electrodes and benchtop control unit(s) (Figure 1A-B). Each electrode is 12mm in diameter, spaced 25mm apart, and densely cover the forearm from elbow to wrist. In this study, the sleeve was donned and doffed by a zipper along the ulnar edge of the garment. Prior to donning, a conduction enhancing spray (Signaspray, Parker Laboratories, Inc.) was applied to the forearm and then the sleeve was placed on the forearm. A custom mixed ionic-electronic conductor (MIEC) enhancer sheet was placed between the electrodes and the skin to improve comfort during stimulation^30^.

**Figure 1.**
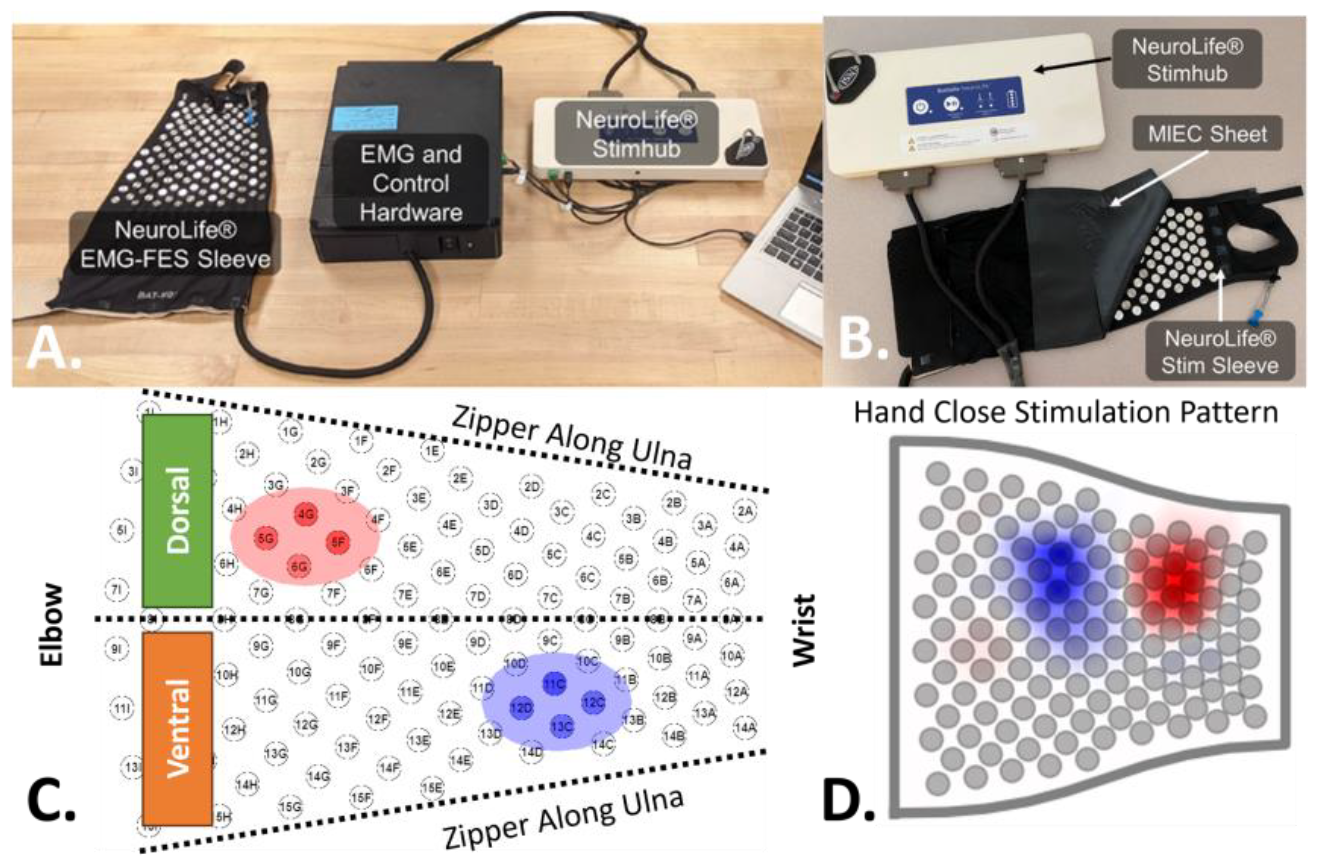
NeuroLife Sleeve systems. A) EMG-controlled FES system B) Operator-controlled FES system stimulator (NeuroLife StimHub) and sleeve with conduction enhancer sheet (MIEC sheet) C) FES pattern calibration user interface with draggable and resizable anode and cathode ellipses. D) Average Hand Close pattern configuration for subject 2, blue indicates cathodic current amplitude, red indicates anodic current amplitude.

Stimulation configurations for each movement (FES patterns) were calibrated through a software graphical user interface that allows for the independent selection of anode and cathode electrodes, stimulation frequency (20-50Hz), and stimulation amplitude (up to 20mA per electrode) (Fig. 1C). Stimulation amplitude was spatially modulated across the ellipses by a Gaussian-like function with tunable drop-off from center. All other stimulation parameters were kept constant, and asymmetric pulses were delivered (Phase 1: 500μs, Phase 2: 1000μs, inter-pulse duration: 200μs). Before the first therapy session, each subject underwent an initial calibration process to identify anode and cathode locations and amplitudes for each target movement. Stimulation frequencies were tailored to each movement in operator-controlled sessions but were fixed to 20Hz in EMG-controlled sessions to facilitate EMG decoding. The combination of active electrodes and stimulation parameters are then fixed for each movement which together we term a FES pattern. At subsequent sessions, the previous FES pattern files were loaded and adjustments to the active electrode locations and current amplitude were made to account for day-to-day shifts in sleeve placement, muscle strength and coordination.

In the operator-controlled FES sessions, the FES-only version of the NeuroLife Sleeve system (Figure 1B) was used. In these sessions, participants were instructed to attempt functional tasks normally, and the operator anticipated the subject’s intention and manually activated appropriate FES patterns via GUI buttons to assist with movement. In the EMG-controlled FES sessions, the EMG-controlled FES system (Figure 1A) was used, which has bidirectional electrodes capable of acquiring EMG between stimulation pulses. EMG signals in the EMG-controlled FES system were acquired at 3kHz sampling rate with a gain of 192 V/V using Intan Electrophysiology Amplifiers (Intan RHD2000, Intan Technologies, Los Angeles, CA)^31^. Decoding algorithms determined the intended movement (or rest) from EMG signals and triggered the appropriate FES pattern in real time.

### Decoding Algorithms

To enable participant’s volitional control of the FES, real-time classification algorithms were trained at each session using EMG signals to predict intended movements. Several minutes of data, broken into cued “blocks”, were collected at the beginning of each session for algorithm training. In each block, subjects were visually cued to perform isolated movements or to execute a functional task in steps. To label the functional task data, at most two functional movements were identified as components for each task, and labels were assigned to the component task step (e.g. Hand Open was labeled during the reaching phase and Hand Close was labeled during the grasp phase of the ball grasping task in 2C). These blocks were repeated with and without assistive FES delivered. EMG data were bandpass filtered (20-400Hz, 10^th^ order Butterworth filter) with a 60Hz notch filter. The root mean square (RMS) values for each channel were computed for 100ms non-overlapping bins and a 600-ms shift in cue labels was applied to account for the reaction time of the subject (Figure 2A). Neural network classifier models were trained on samples of four consecutive RMS bins and component movement labels for each task to be practiced in a session. The model input was a flattened array of bins and channels connected to two densely connected layers of 1000 and 500 nodes. A Softmax activation function was applied to the model outputs to provide prediction probability for each movement. (Figure 2B). A full description of the model can be found in the supplementary material. To test the performance of the decoding algorithms, an additional block of cued tasks was collected. If the computed bin-wise accuracy for the test block was below 80%, the classifier was iteratively retrained by adding the previous test block to the training set and collecting a subsequent test block. Complete description of the real-time algorithms can be found in our previous publication^32^.

**Figure 2.**
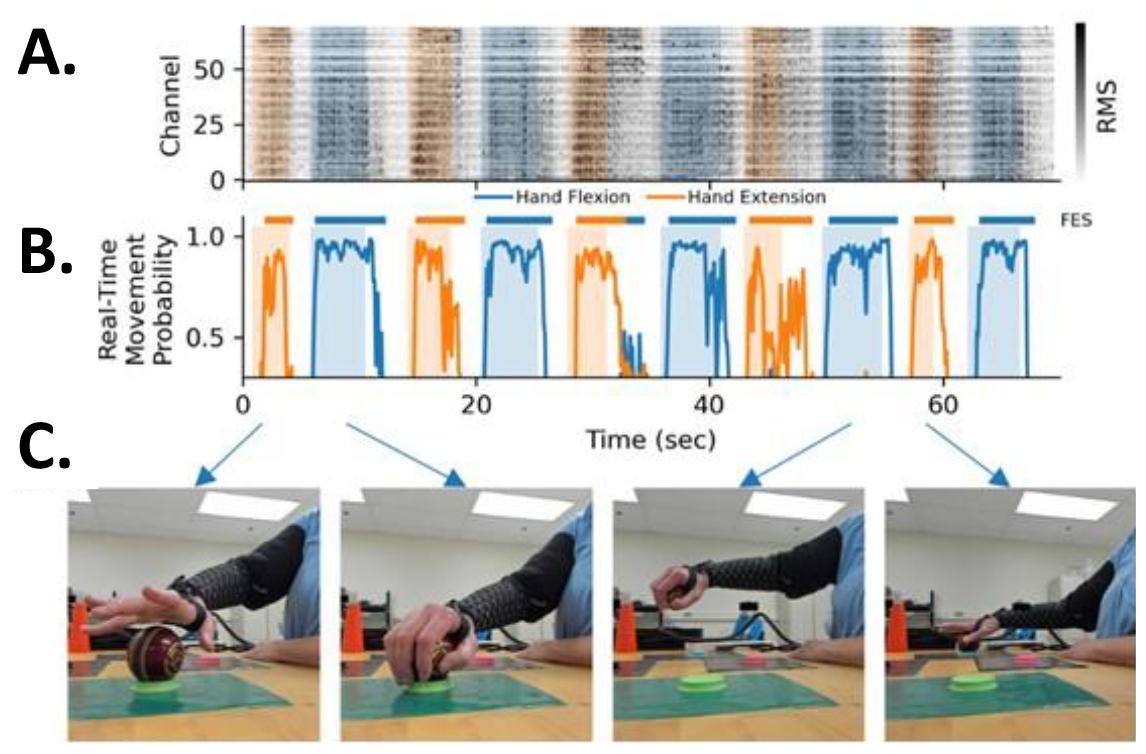
NeuroLife Sleeve operation. A) RMS EMG amplitude over the 70 EMG channels. Shading indicates the movement observed by the operator. B) Movement decoder probability. Bars over the plot indicate FES delivery based on decoder prediction. C) FES-enabled movement used for afunctional task (transferring a ball).

**Figure 3.**
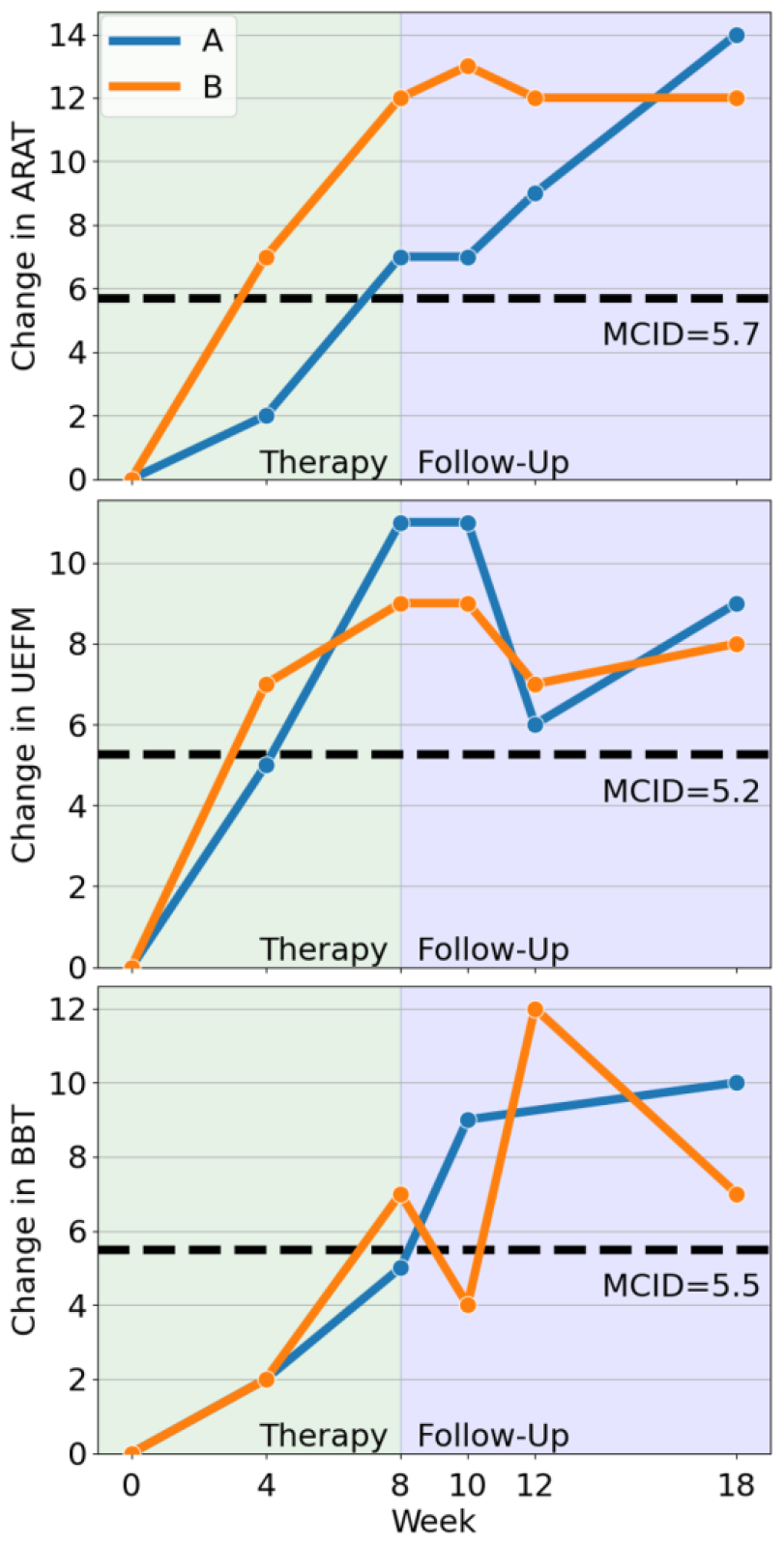
Change in motor recovery at the outcome timepoints. A) Action Arm Research Test (ARAT). B) Upper Extremity Fugl Meyer (UEFM). C) Box and Blocks Test (BBT). Black lines indicate the minimal clinically important difference (MCID). Green shading indicates the therapy intervention period, and blue shading indicates the follow-up period.

### Experimental Setup

During the 8-week intervention period, therapy was administered during three 2-hour sessions with the operator-controlled system used twice per week, and the EMG-controlled system used once per week. In these sessions all of the subject’s FES patterns were recalibrated from previously created patterns. In operator-controlled sessions, an operator matched FES to the subject’s intention during occupational therapist (OT)-guided therapy. At each session, the OT chose functional tasks, graded them to an appropriate difficulty, and administered practice for approximately 20 minutes before moving on to a new task. Subjects were allowed time to rest between tasks and when requested to prevent fatigue. In EMG-controlled sessions, FES patterns were recalibrated and movement intention decoders were trained before therapy. Only tasks using hand open and close FES-enabled movements were used in EMG-controlled sessions. TOT was administered by researchers trained by the OT in this study. The subject’s arm was inspected before donning and after doffing the sleeve to ensure there were no observable changes to the skin.

Subjects were monitored in a 10-week follow-up period during which TOT was not administered. Passive FES and intention-driven FES was delivered during this period to enable continued engineering development of the NeuroLife Sleeve systems. FES during this time was not provided in regularly scheduled sessions and was not directed towards providing therapeutic intervention. Instead, the goal was to collect data for advancement of EMG decoding algorithms and reliability of FES pattern calibration.

### Clinical Assessments

Clinical assessments were performed at the following timepoints: before the intervention (Pre), 4 weeks during intervention (Mid), 8 weeks immediately at the conclusion of the intervention (End), 2 weeks post-intervention (Post-2), 4 weeks post-intervention (Post-4), and 10 weeks post-intervention (Post-10). Clinical assessments included the Action Research Arm Test (ARAT), Upper Extremity Fugl Meyer (UEFM), and the Box and Blocks Test (BBT), Nine-Hole Peg Test (NHPT). Time limits were not imposed on the NHPT assessment, but subjects were allowed to discontinue the test if they felt unable to complete it. The Motor Activity Log (MAL) was completed at the end of operator-controlled sessions as a means to discuss strategies for incorporating hand use in real-world settings as part of the Transfer Package method^33^. These assessments were chosen to span the domains of the World Health Organization International Classification of Functioning (ICF) framework in order to assess the holistic changes during and after intervention^34^. This study was registered on ClinicalTrials.gov (NCT06207240).

## Results

Two adults with upper extremity hemiparesis due to stroke were enrolled in the study. These subjects were both over 60 years of age and in the chronic phase of stroke recovery (> 2 years). Impairment of both subjects was classified as moderate at baseline assessment based on UEFM scores^35^ (Moderate: 19±2 to 47±2 points, A: 28 points, B: 34 points). Data were collected as part of an ongoing clinical study being conducted at Battelle Memorial Institute that was approved by the Battelle Memorial Institute Institutional Review Board (IRB0828, IRB0779). All subjects provided written informed consent before participation, in accordance with the Declaration of Helsinki. No serious unexpected adverse events nor device-related adverse events occurred during the study intervention or follow-ups.

### NeuroLife system enabled intention-driven FES with a variety of tasks

Descriptive statistics of therapy are reported as median (inter-quartile range) for each subject. Subject A received an average of 145 (95-174) stimulations per session with an average therapy duration of 51.8 (44.0-74-7) minutes in each session, similar to previous FEST studies^17,36^. Subject B received an average of 156 (123-189) stimulations per session with an average therapy duration of 49.5 (42.0-70.7) minutes in each session. Each intention-driven stimulation event lasted an average 2.0 (1.27-3.20) seconds for subject A and 1.86 (1.06-3.05) seconds for subject B. Over the course of the intervention, 16 task-oriented therapy activities were practiced with each subject. FES patterns were created to assist with 6 movements for subject A and 5 movements for subject B. An average of 2 (2-3) movements were used per session for both subjects in operator-controlled sessions. EMG-controlled sessions were restricted to Hand Open and Hand Close movements, both of which were used in every session.

### Intention-driven FES with task-oriented therapy improved upper limb function

At the conclusion of the 8-week therapy schedule, both subjects demonstrated improvements that exceeded the minimal clinically important difference (MCID) in at least two assessments^37–39^ (Table 1, Figure 4). ARAT scores improved beyond the MCID (A: +7, B: +12, MCID: +5.7). UEFM scores improved beyond the MCID (A: +11, B: +9, MCID: +5.25). BBT scores improved for both subjects and beyond the MCID for subject B (A: +5, B +7, MCID: +5.5). 9HPT scores improved to 158 seconds for subject A and 53.9 seconds for subject B, whereas both subjects were unable to perform the test at the pre-intervention timepoint (Pre). Although the MAL was not an outcome measure for this study, the MAL scores improved above MCID for both Amount of Use (A: +1.69, B: +1.78, MCID: +1.00) and Quality of Movement (A: +1.92, B: +1.75, MCID: +1.10). The BBT for subject A was not administered at the 4-wk follow-up timepoint due to an injury that was determined unrelated to the study.

**Table 1.** Summary of clinical assessment scores at all timepoints.

| Assessment | Subject | Pre<br>0 | Mid<br>4 | Post<br>8 | Post-2<br>10 | Post-4<br>12 | Post-10<br>18 |
| --- | --- | --- | --- | --- | --- | --- | --- |
| **ARAT**<br>**MCID: 5.7** | A | 25 | 27 (+2) | 32 (+7)* | 32 (+7)* | 34 (+9)* | 39 (+11)* |
|  | B | 35 | 42 (+7)* | 47 (+12)* | 48 (+13)* | 47 (+12)* | 47 (+12)* |
| **UEFM**<br>**MCID: 5.25** | A | 28 | 33 (+5) | 39 (+11)* | 39 (+11)* | 34 (+6)* | 37 (+9)* |
|  | B | 34 | 41 (+7)* | 43 (+9)* | 43 (+9)* | 41 (+7)* | 42 (+8)* |
| **BBT**<br>**MCID: 5.5** | A | 13 | 15 (+2) | 18 (+5) | 22 (+9)* | - | 23 (+10)* |
|  | B | 28 | 30 (+2) | 35 (+7)* | 32 (+4) | 40 (+12)* | 35 (+7)* |
| **NHPT** | A | Unable | 234 | 158 | 105 | 116 | 97 |
|  | B | Unable | 78 | 53.9 | 51.6 | 70 | 91 |
| **MAL - AOU**<br>**MCID: 1.0** | A | 0.77 | 1.85<br>(+1.08)* | 2.69<br>(+1.92)* | - | - | - |
|  | B | 1.35 | 2.35<br>(+1.00)* | 3.13<br>(+1.78)* | - | - | - |
| **MAL - QOM**<br>**MCID: 1.1** | A | 0.85 | 1.96<br>(+1.11)* | 2.54<br>(+1.69)* | - | - | - |
|  | B | 1.21 | 2.38<br>(+1.17)* | 2.96<br>(+1.75)* | - | - | - |

Assessment scores and change from Pre timepoint in parentheses. Changes exceeding the MCID are indicated by (\*). Assessments not administered are indicated by (-). Missing entries were not possible to compute.

### Recovery persisted after intervention with significant dose reduction

The therapeutic improvements observed during the first 8 weeks were sustained during the 10-week follow-up period. During this time, subjects continued receiving FES at a reduced dose, constituting an 84.4% reduction for subject A and 96.4% reduction for subject B in mean weekly stimulation duration (see Supplementary Fig. 2). The ARAT and UEFM improvements remained clinically meaningful for both subjects during the 10 weeks following intervention (Figure 4). Subject A demonstrated continued improvement on the ARAT assessment during the follow-up period.

## Discussion

In this study, two subjects underwent 8 weeks of intention-driven FEST 3 times per week. At each session, subjects received approximately one hour of FES-enabled practice with tasks using multiple functional movements. Both subjects showed clinically meaningful functional improvements throughout the intervention period, as shown by improvements in the ARAT, UEFM, NHPT, and BBT assessments. Improvements persisted through the 10-week follow-up despite greatly reduced FES dose. The session duration and number of repetitions per session in this study were greater than those observed in the clinic with an equal session frequency^40^. These results provide an initial demonstration of the NeuroLife Sleeve’s capability to administer intention-driven FEST incorporating multiple tasks and movements to support functional recovery in chronic stroke survivors.

These results follow previous research on the effects of FEST on neuroplasticity and functional recovery. The intervention in this case study comports with recommendations for FEST administration: mandatory volitional effort from participants, electrical stimulation evoking functional task-related movement, and therapist guidance during task execution^11,14^. The duration (8 weeks) and weekly dose (∼1 hour, 3 times per week) of this intervention are also similar previous studies of FEST for upper limb stroke rehabilitation^12,14,17,36^. Previous studies of FEST for upper limb stroke rehabilitation have shown clinically meaningful improvements to upper limb function, such as shown in this case series^12,13,23,29^.

Pairing FES with motor intention is proposed to engage neuroplastic mechanisms of recovery. In case studies of EEG-controlled FEST with comparable durations and upper-limb tasks, similar improvements have been observed in ARAT and UEFM scores in chronic stroke survivors^17,36^. In these studies, EEG was used to initiate a timed sequence of FES patterns, whereas in the FEST sessions here, operators and EMG decoders were used to control both the initiation and termination movements continuously to execute functional tasks. Jonsdottir et al. (2017) observed improvements to ARAT and UEFM scores, similar in magnitude to those reported here, in their chronic study subjects using the Myoelectric Controlled FES (MeCFES) system enabling continuous and proportional control of FEST^23,28^. This enhanced control may provide a greater sense of agency to the user and more consistent active participation, enhancing the neuroplastic and functional benefits of the therapy.

However, EMG-controlled FES has not shown clear superiority to other interventions in meta-review^22^. This may be due to sub-optimal, inconsistent parameters and hardware limitations to use cases used to deliver EMG-controlled FES. Another inconsistency in EMG-controlled and intention-driven FES upper limb rehabilitation literature is the number of muscles, movements, and tasks targeted in interventional studies. Two recent randomized clinical trials showed improvements to upper extremity function with EMG-controlled FES practice of wrist and finger extension that were non-significant compared to cyclic FES (Wilson et al.)^20^ or conventional therapy alone (Kwakkel et al.)^41^. Taken with the significantly greater improvements compared to conventional therapy found by Thorsen et al.^42^ may indicate that EMG-controlled FES is better applied for TOT than isolated movement practice alone.

Furthermore, more limited response to EMG-controlled FES interventions has been observed in subjects with higher baseline upper limb impairment^22,23,41^. EMG control schemes require some volitional muscle activation remaining after injury to control FEST, which may make EEG control more feasible for more impaired users^17^. Kwakkel et al. and Jonsdottir et al. note that EMG-controlled FES interventions were adapted for subjects with EMG signals insufficient for FES control by placing EMG electrodes on contralateral unaffected or synergistic muscles^23,41^. While these enable therapy, reduced coupling of motor intention and FES sensory activation in this off-target control scheme may be suboptimal to drive neuroplastic effects. Subjects in the present case series were both categorized as moderately impaired (UEFM between 19 and 47 points) and in the chronic (>9 months post-stroke) at baseline^35^. Our group has previously shown continuous EMG-based decoding of multiple movements using the NeuroLife Sleeve in severe chronic stroke survivors (UEFM hand subscore <3)^32^. However, further studies are needed to determine whether similar results are possible with the additional complexity of decoding through stimulation, which allows for recording and stimulation through the same electrodes in this system.

Finally, to facilitate integration with existing clinical practices, an intention-driven FEST system should readily allow for adjustment to task challenge and type throughout the session^26,43^. In the *Practical Considerations for Therapist* section of Kapadia et al.’s review of FEST for reaching and grasping, 3 of 12 general procedure steps concern placement and calibration of patch FES electrodes, highlighting the complexity and import of this aspect of setup^14^. The NeuroLife Sleeve is designed for consistent placement of electrodes via zipper closure aligned from ulnar styloid process to elbow. Combined with the ability to quickly change active electrodes in the array and save and load previous FES patterns in the GUI, this system reduces the manual burden of electrode placement and recalibration.

While results from this case series are encouraging, several limitations warrant discussion. The small sample size (N=2) and lack of control group in this study prevent any statistically driven conclusions about this intervention. Future work will include a randomized controlled clinical trial to directly compare the effects of intentional control using the sleeve against timer-based FES. Two different methods were used to estimate motor intention and control of the FES during this study. Operator control was provided in two out of three weekly sessions while EMG control was provided in the third. Because both control methods were employed throughout the study, we cannot distinguish between the therapeutic effects of each. In future studies, only EMG control will be used to reduce the operational cost of the system. EMG-controlled FES sessions were limited to tasks using gross movements (hand open and hand close) to ensure robust decoder performance and intentional control. Further refinement of our decoding algorithms will enable higher degrees of freedom and more robust detection of fine motor control. Though the NeuroLife Sleeve system provides advantages over other FES and intention-driven FES systems, an experienced operator was required at all therapy sessions in this study to ensure proper calibration of FES patterns and decoder algorithms. Future development will focus on designing a therapist interface, reducing the hardware footprint, and reducing calibration time to increase the therapy dose at each session.

Additional studies will be needed to establish the safety, feasibility, and efficacy in a larger stroke survivor population. The upwards slope between the mid- and post-intervention assessments suggests that further improvement could be possible with a longer course of therapy, motivating studies of optimal therapy dosing. The benefits from this intervention remained greater than the MCID at least 10 weeks after the end of therapy, though participants continued to receive a low dose of FES. This indicates that this intervention may have driven neuroplasticity that led to persistent functional improvements and that these improvements could be maintained with a drastically reduced dose. However, the increase in volitional daily use of the affected arm, evidenced by the increased MAL scores, likely contributed to the functional improvement in the intervention period and may have driven the post-intervention improvement in subject A (Figure 4). Anecdotally, both subjects reported personally meaningful functional recovery over the course of this study. Subject A self-reported that they were able to tie their shoes for the first time since their stroke more than 4 years earlier. Subject B self-reported they started spontaneously using their paretic hand while preparing dinner, which was their dominant hand before their stroke.

## Conclusion

In this case series we present two chronic stroke subjects with improved upper limb function following 8 weeks of (1) FES during task-oriented therapy (2) controlled by user intention (3) spanning multiple movements. Intention-driven FES was delivered using the NeuroLife Sleeve system, a wearable sleeve with an embedded high-density electrode array. Outcome measures showed clinically meaningful improvements (above the MCID) over the intervention period, during which subjects received 1 hour of therapy 3 days per week. Furthermore, clinically meaningful improvements were sustained for most assessments above MCID throughout the 10-week follow-up period with a low dose of FES. The intervention integrated well with standard clinical practice for task-oriented therapy. This study provides evidence intention-driven FEST interventions can facilitate clinically meaningful change using wearable FES technology to enhance usability and task flexibility.

## Data Availability

All data produced in the present study are available upon reasonable request to the authors

## Disclaimer

These devices have not been approved or cleared as safe or effective by FDA. These devices are limited by U.S. federal law to investigational use.

