## Supplementary Materials for "Restoration of upper-extremity function after task-oriented, intention-driven functional electrical stimulation therapy using a wearable sleeve in adults with chronic stroke: a case series"

### Supplementary Methods

#### IRB0828 Study Inclusion and Exclusion Criteria

Inclusion Criteria

1) Males and females ≥18 years old
2) Diagnosis of stroke
3) Ability to provide appropriate consent to partake in the study
4) Ability to follow 3-step commands and deemed by an occupational therapist to have the capacity to complete required upper extremity movements
5) Ability to secure transportation to attend scheduled study sessions
6) Stroke-related hand impairment that interferes with ability to complete activities of daily living and is classified as Stage 1-6 on the hand subscale of the Chedoke McMaster Stroke Assessment

Exclusion Criteria

1) Presence of any other clinically significant medical comorbidity for which, in the judgment of the Investigator, participation in the study would pose a safety risk to the subject
2) Currently participating in physical rehabilitation (e.g., occupational or physical therapy) for stroke-related upper limb impairment
3) Co-occurring neurological condition (e.g., Parkinson’s disease, Multiple Sclerosis) or other neuromuscular disorder (e.g., Carpal Tunnel Syndrome, neuropathy) that, in the judgment of the Investigator, may influence study results
4) Individuals who are immunosuppressed, have conditions that typically result in becoming immunocompromised, taking chronic steroids, or currently receiving immunosuppressive therapy
5) Individuals having or requiring any of the following: implanted pacemaker, life supporting/sustaining equipment, or critical non-removable implantable electronic devices such as an insulin pump.
6) Persistent pain ≥ 7/10 in impaired upper extremity, as measured by Visual Analogue Scale
7) Individuals whose forearm is determined to be too small or too large to fit the electrode sleeve being investigated.
8) Individuals who are pregnant or plan to get pregnant during the course of the study (self report).

#### Follow-up Period

Clinical assessments were administered at three follow-up timepoints after the end of the intervention period. During the follow-up period, both subjects received FES as part of a study supporting the development of the EMG-FES NeuroLife Sleeve system (IRB0779). Sessions in this study were scheduled ad hoc and less frequently than those in the study presented in the manuscript. These sessions were not designed to provide therapeutic intervention but included elements that have been shown to benefit stroke survivors. Details of FES dose in the follow-up period are presented here (Supplemental Figure 4).

#### Decoding Algorithm Model

### The NN was developed in Python 3.8 using the FastAI package [1]. FastAI defaults were used for training except where noted. The model architecture takes an input of a flattened N channels x 4 array from the N channels of the sleeve and 4, 100ms windows of mean RMS signal. The input layer connects to two fully-connected dense layers, with size 1000 and 500 respectively, with batch normalization and the ReLU activation function between layers. The final layer had 13 classes corresponding to the 12 cued movements and rest. Finally, a Softmax activation function was applied to the model outputs to provide prediction probabilities for each of the movements. The predicted movement for a given time point was the movement with the greatest prediction probability. The training procedure used label smoothing cross entropy loss (p=0.9) and the Adam optimizer. During training, dropout was applied to each layer with 20% probability to prevent overfitting. The learning rate was optimized using the FastAI learning rate finder tool [1]. Each model was trained for 400 epochs with early stopping criterion, using the one cycle training policy from FastAI.

1. Howard J and Gugger S 2020 Fastai: A Layered API for Deep Learning Information 11

### Supplemental Figures

#### 1. Number of FES-Enabled Stimulations

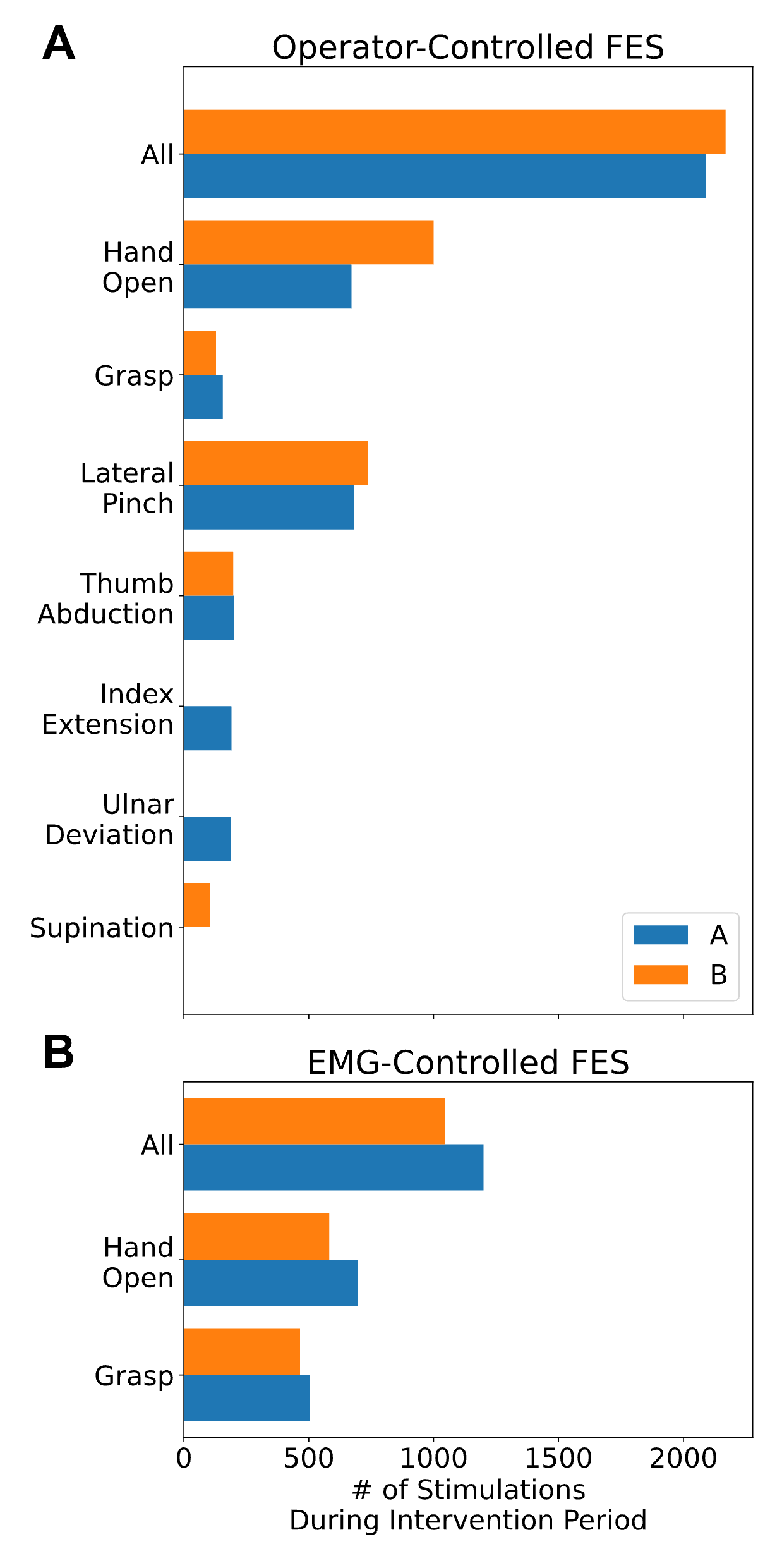

Breakdown of stimulations by movement delivered over 8 weeks of intention-driven FES therapy. A) Total number of operator-controlled stimulations. B) Total number of EMG-controlled stimulations.

#### 2. FES-Enabled Movements in Functional Tasks

| Task-Oriented Activity | FES-enabled Movements | Subjects | Description |
| --- | --- | --- | --- |
| Cone Stacking | Hand Open, Thumb abduction, Grasp | A, B | Stacking and unstacking cones 8” cones. |
| Cup Pour | Hand Open, Grasp, Supination | A, B | Grasping cup from upright position then pronating to pour, and reverse to upright. |
| Coin Stacking | Lateral Pinch | A, B | Picking up coins from table and stacking them. |
| Ball Gathering | Lateral Pinch, Thumb abduction | A, B | Gathering small balls from table into cup receptacle. |
| Cutting Playdoh | Grasp, Lateral Pinch | A, B | Cutting flattened playdoh with butter knife. |
| Shut the Box | Index Extension | A, B | Turning down tiles from the game “Shut the Box” (<https://www.mastersofgames.com/cat/pub/shut-the-box.htm>) |
| Removing Playdoh Cap | Lateral Pinch | A, B | Removing the cap from a playdoh container. |
| Connect Four | Lateral Pinch | A, B | Picking up and placing Connect Four tiles |
| Clothespins | Lateral Pinch, Thumb abduction | A, B | Clipping clothespins on a string. |
| Wiping surface | Ulnar deviation | A | Sliding a towel on the surface of a desk using only wrist rotation. |
| Opening pill bottle | Hand Open | A, B | Opening child-safe pill bottle |
| Playdoh Squeeze | Grasp | A, B | Repeated strong squeeze of Playdoh. |
| Playdoh Pinch | Lateral Pinch | A, B | Removing small objects embedded in Playdoh with thumb. |
| Thumb to finger tip | Thumb abduction | B | Touching each finger tip to thumb consecutively. |
| Playdoh meal simulation | Lateral pinch | A, B | Using fork to skewer pieces of playdoh and raise to mouth. |
| Salad tongs | Lateral pinch | B | Opening and closing tongs and supinating/pronating to place imaginary salad. |
| Keyboard typing | Index extension | A | Typing on a keyboard or cellphone with index finger. |
| Ball Transfer | Hand open, Grasp | A, B | Grasping a cricket ball, moving it to a new location, and releasing. |
| Cone Transfer | Hand open, Grasp | A, B | Grasping an 8” cone, moving it to a new location, and releasing. |

Task-oriented therapy activities used in the intervention period. **Task-oriented activity:** Task short name, **FES-enabled Movement:** the movements used to assist the task, **Subjects:** which subjects were given the task during the intervention, **Description:** brief description of the activity. Note that the objects used and exact activity procedure was varied to grade the task difficulty, but the description is representative of the activities. Not all of the listed FES-enabled movements were used at every instance of activity practice (e.g. sometimes stimulation was only paired with Hand Open intention, not Thumb Abduction or Grasp). The FES-enabled movements used in the task were chosen based on which were most helpful to the subject in that session.

#### 3. Intervention and Follow-Up FES Dose

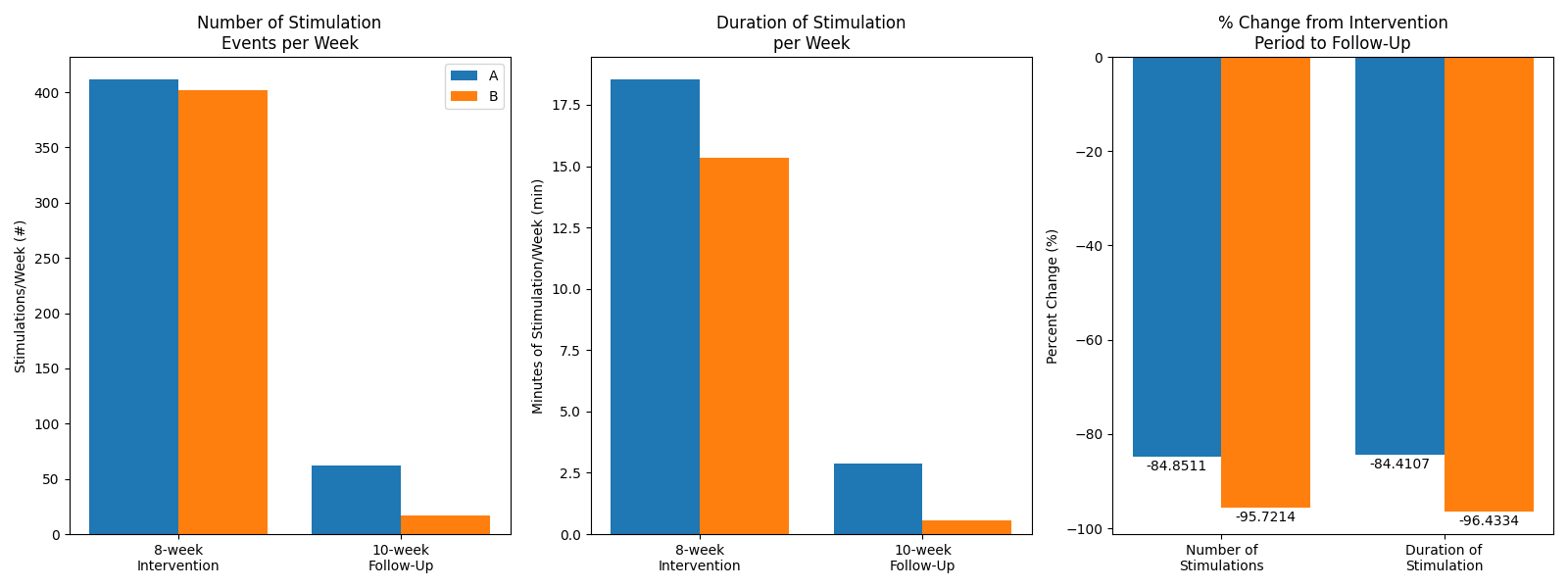

The number of stimulation events (left) and the duration of stimulation (middle) delivered weekly decreased for both subjects from the intervention period to the follow-up. This constituted an 84% reduction for subject A and a 96% reduction for subject B in duration of weekly stimulation (right).

#### 4. FES Patterns Calibrated Using the NeuroLife Sleeve

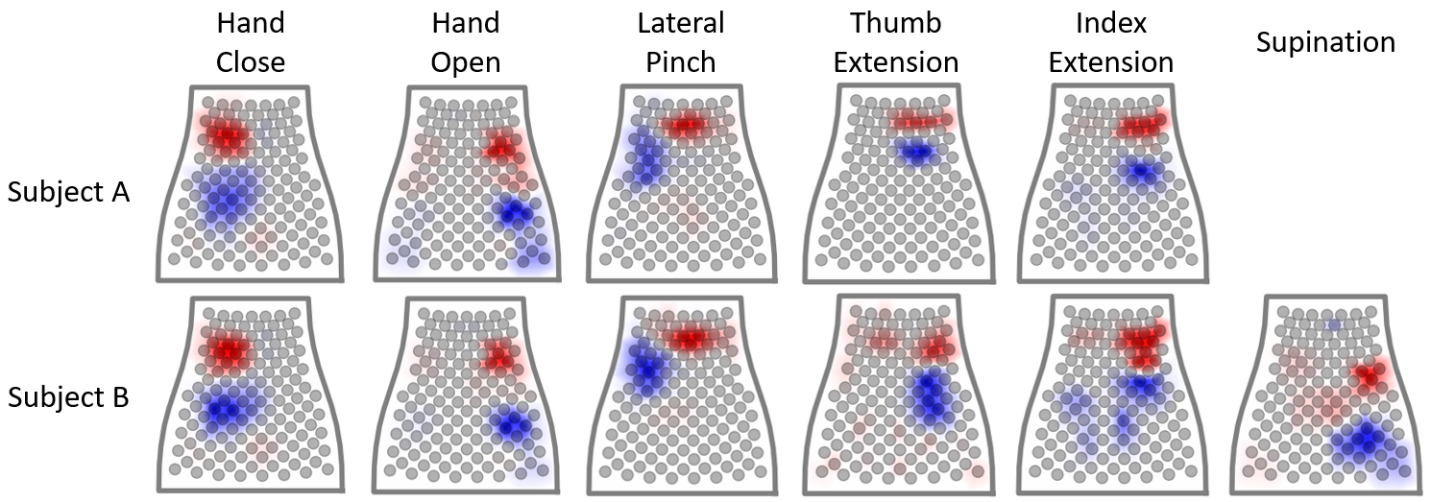

Active electrodes in the high-density array for calibrated FES patterns averaged over all sessions. FES patterns for 5 movements were calibrated for Subject A, and 6 for Subject B.
